# Prevalence of use and interest in using glucagon-like peptide-1 receptor agonists for weight loss: a population study in Great Britain

**DOI:** 10.1101/2025.06.06.25329114

**Authors:** Sarah E. Jackson, Jamie Brown, Clare Llewellyn, Oliver Mytton, Lion Shahab

**Affiliations:** Department of Behavioural Science and Health, University College London, UK; Institute for Child Health, University College London, UK

**Author notes:** Corresponding author: Dr Sarah Jackson, Department of Behavioural Science and Health, University College London, 1-19 Torrington Place, London WC1E 7HB, UK.

**Keywords:** semaglutide, liraglutide, tirzepatide, Ozempic, Wegovy, Saxenda, Mounjaro

## Abstract

**Objectives:** To assess the prevalence of glucagon-like peptide-1 receptor agonist (GLP-1RA) use and interest in using medications for weight loss among adults in Great Britain.

**Design:** Nationally-representative household survey, January-March 2025.

**Setting:** Great Britain.

**Participants:** 5,893 adults (≥18y).

**Main outcome measures:** Participants were asked whether they had used medication in the past year to manage type 2 diabetes (excluding insulin), reduce the risk of heart disease, or support weight loss and, if so, whether they had used five specific GLP1-RAs. Those who had not used medication to support weight loss in the past year were asked how likely they would be to consider doing so in the next year. Estimates were reported stratified by participant characteristics and extrapolated to the national population.

**Results:** Overall, 2.9% [2.4-3.4%] – ∼1.6 million adults – reported using a GLP-1RA to support weight loss in the past year, with 1.7% [1.4-2.1%] (∼910,000 adults) using them exclusively for this purpose. The majority of those who used them exclusively for weight loss (91.4% [85.6-97.2%]) reported using GLP-1RAs that are licensed for this purpose in Great Britain, most commonly Mounjaro (tirzepatide; 80.2% [71.9-88.6%]). Of those who had not used weight-loss medication in the past year, 6.5% [5.7-7.3%] (∼3.3 million adults) expressed an interest in doing so in the next year. Use and interest were more prevalent among women, people in mid-life, and those reporting past-month psychological distress. Interest was also higher among people facing greater socioeconomic disadvantage, including those in financial difficulty or unemployed due to long-term illness or disability.

**Conclusions:** In the first quarter of 2025, 4.2 million adults in Great Britain – nearly one in ten – either had recently used a GLP-1RA to support weight loss or were interested in doing so in the near future. A substantial minority reported using a type of GLP-1RA that was not licensed for weight management, suggesting off-label use. Interest was particularly high among less advantaged socioeconomic groups, while use was similar across groups, highlighting the importance of addressing equity in access. These findings underscore the need to monitor who is accessing these medications and to ensure their safe, appropriate, and equitable provision.

**Registration:** The study protocol and analysis plan were pre-registered on Open Science Framework (https://osf.io/r2whq/).

## Introduction

The high prevalence of obesity in Great Britain presents significant public health challenges, increasing the risk of chronic diseases and placing a substantial burden on the National Health Service (NHS).^1,2^ Advancements in pharmacotherapy, particularly glucagon-like peptide-1 receptor agonists (GLP-1RAs), have emerged as a promising tool for weight management. Originally developed and increasingly used for the treatment of type 2 diabetes, these medications have proven highly effective in promoting weight loss, at least during treatment,^3–5^ sparking widespread public interest. They also have cardio-protective effects, reducing the risks of heart attack, stroke, and cardiovascular mortality^6,7^ and may help to reduce substance use disorders (e.g., alcohol and tobacco use),^8^ but safety of long-term use is currently uncertain.^9^ Relatively little is known about the current prevalence of the use of GLP-1RA medications in Great Britain,^10^ potential future demand, and how interest and usage patterns vary across different population subgroups. Understanding these factors is important for informing healthcare planning and resource allocation, promoting patient safety, and addressing health inequalities.^11^

In the UK, several GLP-1RA medications are available, including semaglutide, liraglutide, dulaglutide, exenatide, and lixisenatide. These are supplied under various brand names, some of which have more than one indication (see ^12^ for a summary). In addition, tirzepatide is a GLP-1RA combined with glucose-dependent insulinotropic polypeptide receptor agonist. GLP-1RA products currently licensed for weight loss in the UK – Saxenda (liraglutide), Wegovy (semaglutide), and Mounjaro (tirzepatide) – are licensed by the Medicines and Healthcare products Regulatory Agency for patients who (i) have obesity (body mass index [BMI] ≥30 kg/m²) or (ii) have a BMI in the overweight range (BMI ≥27 kg/m²) plus weight-related comorbidities, such as cardiovascular disease.^12^ Clinical guidelines from the National Institute for Health and Care Excellence (NICE),^13^ making recommendations to the NHS to ensure value for money, set higher threshold for use.

NICE recommend tirzepatide and semaglutide for patients with at least one weight-related comorbidity and a BMI ≥35 kg/m² (or BMI ≥30 kg/m² and who meet the criteria for referral to specialist weight management services, for semaglutide only) and liraglutide for patients with a BMI ≥35 kg/m², non-diabetic hyperglycaemia, and a high risk of cardiovascular disease. In England this would equate to 3.4 million being eligible for the medications on the NHS.^14^ Given concerns about cost and resources pressure on the NHS, the NHS in England plans a phased roll out, offering the drugs to 220,000 people between 2025 and 2028.^14^

Public interest in, and use of, GLP-1RAs for weight loss is growing rapidly. In the United States, a poll conducted in May 2024 (n=1,479) found that 6% of adults were currently using a GLP-1RA and 12% had ever used one.^15^ Among those who reported ever taking the drugs, most (61%) said they took them to manage a chronic condition such as diabetes or heart disease, but around four in ten (38%) said they took them solely to lose weight.^15^ Similarly, demand for these medications in the UK appears to be high, with concern that the NHS is struggling to manage demand for weight-loss drugs.^16^ Prescribing data show sharp increases in prescription of semaglutide and tirzepatide across NHS GP practices in England in 2024-25.^17,18^ A December 2024 poll (n=2,161) found that 22% of UK adults would use a GLP-1RA weight-loss drug if it were available on prescription through the NHS,^19^ although many of these may not be eligible according to NICE guidelines. Of those surveyed, 5% had personally taken a GLP-1RA, while 9% knew friends or family members who had used one.^19^

Despite the growing interest in these medications, there is also concern about their misuse. Anecdotal evidence and adverse drug reaction reports suggest some people are using GLP-1RAs outside of licensed indications,^12^ potentially posing health risks. Additionally, gastrointestinal side effects (e.g., nausea, vomiting, diarrhoea, and constipation) are common,^3,5^ serious adverse effects (e.g. pancreatitis) have been reported,^11^ and some people who use GLP-1RAs experience malnutrition,^20–23^ further raising safety considerations. As these medications become more widely used, it is essential to monitor their usage trends and to understand who is using them. With some people accessing these medications outside of the NHS, traditional systems for understanding usage (e.g., NHS prescribing data, Clinical Practice Research Datalink) cannot be relied upon to provide accurate estimates. Triangulation with nationally-representative surveys is important for providing a complete picture.

Gender differences may play a role in the demand for and usage of GLP-1RA medications. Severe obesity (BMI ≥40) is more prevalent among women than men (4% vs. 2% among adults in England in 2022)^2^ and women are more likely to seek and receive medical treatment for weight loss,^24^ so they may be prescribed these drugs at higher rates than men. Additionally, societal pressures related to body image and weight management disproportionately affect women,^25^ which could further drive interest in GLP-1RAs within this group.

Equity of access to these medications is another important factor.^26^ In the UK poll, just 8% said they would use these medications for weight loss if they had to pay for them privately, compared with the 22% who would take them if provided on prescription by the NHS.^19^ Given it currently costs in the region of £200 per month to obtain these drugs privately,^19^ those with the financial means may have greater access to treatment, potentially exacerbating health inequalities. This is particularly concerning as obesity is more prevalent among socioeconomically disadvantaged groups, particularly among women, according to area level deprivation and household income.^27,28^

Given the rising public interest, potential supply challenges, and safety concerns, this study aimed to assess the current prevalence of GLP-1RA use and level of interest in using these medications for weight loss among the adult population in Great Britain. We also explored differences between key population subgroups. A comprehensive understanding of these factors can help to inform healthcare policy, ensure equitable access to treatment, and safeguard patient wellbeing.

## Methods

### Pre-registration

The study protocol and analysis plan were pre-registered on Open Science Framework (https://osf.io/r2whq/).

### Design

Data were collected via the Smoking Toolkit Study, a monthly cross-sectional survey of a representative sample of adults (≥16 years) in Great Britain (i.e., England, Scotland and Wales). The methods have been described in detail elsewhere.^29–31^ Briefly, the study uses a hybrid of random probability and simple quota sampling to select a new sample of approximately 2,450 adults each month. Data are collected through telephone interviews. Sample weights are calculated using raking to match the population in Great Britain. This profile is determined each month by combining data from the UK Census, the Office for National Statistics mid-year estimates, and the annual National Readership Survey.^29^ Comparisons with other national surveys and sales data indicate the survey achieves nationally representative estimates of key sociodemographic variables.^29^

Between January and March 2025, all participants aged ≥18 years in England and ∼50% in Wales and Scotland were asked additional questions on past-year use of GLP-1RAs and interest in using them for weight loss. This study analysed these data.

### Measures

Past-year use of GLP-1RAs and GLP1-RA compounds (i.e., tirzepatide) was assessed with two questions. The first asked: ‘In the last 12 months have you taken any medication to help you with any of the following, or not?’ Participants were asked to indicate all that applied from the following response options: (a) medication for type 2 diabetes, excluding insulin, to help manage your blood sugar levels; (b) medication to lower the risk of heart disease; (c) medication to support weight loss/ reduce food cravings; (d) I have not taken medication for any of these reasons. They could also respond that they could not remember. Those who reported using medication (i.e., responded a, b, or c) were then asked: ‘In the last 12 months, which, if any, of the following types of medication have you taken [for type 2 diabetes/to lower the risk of heart disease/for weight loss]?’ (a) Saxenda, containing liraglutide; (b) Ozempic, containing semaglutide; (c) Wegovy, containing semaglutide; (d) Mounjaro, containing tirzepatide; (e) Rybelsus, containing semaglutide; (f) another type of medication. Participants were asked to select all that applied. They could also respond that they did not know. Those who responded to one of the named GLP-1RA, i.e. responses (a) to (e), to the second question were considered to have used a GLP-1RA.

Interest in using medication for weight loss was assessed among those who did not report using medication for weight loss in the past year (i.e., responded a, b, or d to the first question) with the question: ‘In the next 12 months, how likely, if at all, are you to consider using a weight loss medication to help you lose weight?’ Response options were: (a) very likely, (b) fairly likely, (c) not very likely, (d) not at all likely, (e) I don’t need to lose weight. Participants could also respond that they did not know. We provided descriptive data for all response options. For some analyses, we dichotomised responses to a or b (likely to consider) vs. all other responses including don’t know.

We captured a range of sociodemographic characteristics. Gender was self-reported as man, woman, or in another way; the latter group was excluded from analyses by gender due to low numbers. Age was analysed as a continuous variable. Ethnicity was categorised as white vs. minority ethnic group. Occupational social grade was categorised using National Readership

Survey classifications^32^ as ABC1 (includes managerial, professional, and upper supervisory occupations) and C2DE (includes manual routine, semi-routine, lower supervisory, state pension, and long-term unemployed). Financial situation was assessed with the question: ‘How well would you say you yourself are managing financially these days? Would you say you are (a) living comfortably; (b) doing alright; (c) just about getting by; (d) finding it quite difficult; or (e) finding it very difficult?’.^33^ Health-related economic inactivity was operationalised as participants reporting that they are ‘not in paid work because of long-term illness or disability’, in response to a question asking which of a list of different working statuses applies to them.

We also recorded information on participants’ health-related behaviours and mental health. Smoking status was categorised as current, former, or never smoker. Level of alcohol consumption was assessed with the Alcohol Use Disorders Identification Test—consumption (AUDIT-C; possible range=0-12) and analysed as a continuous variable. As a general guide, AUDIT-C scores ≥5 indicate drinking at increasing or higher-risk levels (i.e., levels that increase someone’s risk of harm).^34^ Psychological distress was assessed using the Kessler Psychological Distress Scale (K6), which measures non-specific psychological distress in the past month (possible range=0-24).^35,36^ We categorised K6 scores as ‘no/low distress’ (0-4) or ‘moderate/severe distress’ (≥5).^35,37^ History of eating disorders was assessed with the question: ‘Since the age of 16, which of the following, if any, has a doctor or health professional ever told you that you had…?’ Responses were dichotomised to distinguish between those who responded ‘an eating disorder’ and those who did not. Due to limited availability of funding, the question assessing eating disorders was only included in one of the three survey waves (February 2025), so analyses of this variable were restricted to those surveyed in this wave.

### Statistical analysis

Data were analysed using R v.4.4.1. All analyses used weighted data. Missing data were handled using multiple imputation by chained equations. We imputed missing values under the assumption of missing at random, using the *mice* package. Five imputed datasets were generated. The imputation models included all variables used in the analysis (except history of eating disorders, given it was only assessed in one wave; the imputation process was repeated separately for this wave of data to provide complete data for analyses involving this variable). Analyses were conducted separately on each imputed dataset and results were combined across imputations using Rubin’s rules^38^ to account for within- and between-imputation variance. Results were reported as pooled estimates and confidence intervals (CIs).

We estimated proportions (with 95% CIs) of adults who reported having used GLP-1RAs in the past year (a) for any reason, (b) to manage type 2 diabetes, (c) to reduce the risk of heart disease, (d) to support weight loss, and (e) exclusively to support weight loss (i.e., not also to manage type 2 diabetes or to reduce the risk of heart disease). We also reported these prevalence estimates stratified by the specific medication used. Among adults who reported having used a GLP-1RA for weight loss in the past year (at all, and exclusively for weight loss), we estimated the proportion who used a medication licensed for weight loss in Great Britain (Saxenda, Wegovy, or Mounjaro).

Among adults who had not used a medication for weight loss in the past year, we estimated the proportion who would be likely to consider using a weight-loss medication in the next year. We applied these proportions to the most recent (2023) mid-year population estimates for Great Britain^39^ to approximate the number of adults using these medications in 2024 and the number who would be interested in using them in the next year.

To explore differences in (i) past-year use of GLP-1RAs to support weight loss and (ii) interest in using a weight-loss medication in the next year between population subgroups, we estimated these proportions stratified by gender, age, ethnicity, occupational social grade, financial situation, health-related economic inactivity, smoking status, alcohol consumption, past-month distress, and history of eating disorders. We also examined proportions within intersections of gender and each other variable (results are reported in the Supplementary File). Age and level of alcohol consumption were analysed as continuous variables, modelled non-linearly using restricted cubic splines (with three knots placed at the 5, 50, and 95% percentiles) to allow for flexible associations without arbitrary categorisation. Estimates for continuous variables were predicted from unadjusted logistic regression models that tested the association of age/level of alcohol consumption with each outcome. We reported estimates for selected ages (18, 25, 35, 45, 55, 65, and 75) and AUDIT-C scores (0, 3, 5, 8, 11) as an illustrative example of differences across ages and levels of alcohol consumption. We also ran logistic regression models to analyse associations between each participant characteristic and these two outcomes, adjusted for age and gender (or just age, for gender-stratified analyses).

## Results

A total of 5,893 participants were invited to complete the GLP-1RA survey module, of whom 5,260 (89.3%) consented. Characteristics of the total eligible sample and those who consented are shown in **Table S1** alongside imputed data.

### Prevalence of GLP-1RA use

Overall, 4.5% of participants reported using a GLP-1RA in the past year for any reason; 2.9% reported using them to support weight loss, with 1.7% using them exclusively for weight loss (i.e., not also for type 2 diabetes or heart disease; **Table 1**). When extrapolated to the national population, these figures suggest that approximately 1.6 million adults in Great Britain were using GLP-1RAs to support weight loss in early 2025 (53.7 million adults x 2.9%), of whom 910,000 were using them exclusively for weight loss (53.7 million x 1.7%).

**Table 1.** Past-year use of GLP-1 receptor agonists among adults (≥18y) in Great Britain.

| GLP-1RA used in past year | Authorised indication(s)<sup>1</sup> | Prevalence, % [95% CI] |  |  |  |  |
| --- | --- | --- | --- | --- | --- | --- |
|  |  | For any reason | To manage type 2 diabetes | To reduce the risk of heart disease | To support weight loss | Exclusively to support weight loss |
| Any GLP-1RA listed below | - | 4.5 [3.8–5.1] | 1.7 [1.3–2.0] | 1.6 [1.3–2.0] | 2.9 [2.4–3.4] | 1.7 [1.4–2.1] |
| Saxenda, containing liraglutide | WL | 0.4 [0.2–0.5] | 0.2 [0.0–0.3] | 0.1 [0.0–0.3] | 0.1 [0.0–0.2] | 0.0 [0.0–0.0] |
| Wegovy, containing semaglutide | WL, CRR | 0.8 [0.5–1.0] | 0.2 [0.1–0.3] | 0.3 [0.1–0.5] | 0.6 [0.3–0.8] | 0.4 [0.2–0.5] |
| Mounjaro, containing tirzepatide | WL, T2D | 2.3 [1.9–2.7] | 0.6 [0.4–0.9] | 0.7 [0.4–0.9] | 2.0 [1.6–2.4] | 1.4 [1.1–1.7] |
| Ozempic, containing semaglutide | T2D | 1.3 [0.9–1.7] | 0.6 [0.4–0.9] | 0.6 [0.3–0.9] | 0.5 [0.3–0.7] | 0.2 [0.1–0.3] |
| Rybelsus, containing semaglutide | T2D | 0.6 [0.3–0.8] | 0.5 [0.3–0.7] | 0.3 [0.1–0.4] | 0.3 [0.2–0.5] | 0.1 [0.0–0.1] |

CI, confidence interval. GLP-1RA, glucagon-like peptide-1 receptor agonist.
<sup>1</sup> Authorised indications for use in the UK.<sup>12</sup> WL, weight loss. CRR, cardiovascular risk reduction. T2D, type 2 diabetes.
Data shown are weighted estimates of the proportion (with 95% CI) of adults in Great Britain reporting past-year use of different GLP-1RAs, overall (i.e., for any reason) and stratified by the reason for use. Note that reasons are not mutually exclusive, except 'exclusively to support weight loss', which excludes participants reporting use for any other reason.
Estimates stratified by gender are provided in **Table S3**.

There were some subgroup differences in the use of GLP-1RAs to support weight loss (**Table 2**). Prevalence was more than twice as high among women than men (4.0% vs. 1.7%; OR=2.40 [1.62– 3.56]) – with an even greater difference in the proportion using them exclusively for weight loss (2.8% [70.0% of female users] vs. 0.6% [35.3% of male users]; OR=4.44 [2.43–8.15]). There was a non-linear (inverted U-shaped) association with age, with the highest prevalence of use of GLP- 1RAs to support weight loss among those in mid-life (e.g., 4.2% among those aged 45 and 55) and lower prevalence in early adulthood (e.g., 1.2% among those aged 18) and later life (e.g., 1.5% among those aged 75). Prevalence was also higher among those who reported moderate/severe psychological distress (3.7% vs. 2.4% among those reporting no/low distress; OR=1.62 [1.13– 2.32]). It also appeared to be higher among those who reported a history of eating disorders (4.4% vs. 2.1% among those who did not; OR=2.12 [0.69–6.53]), but there was substantial imprecision in the estimate for this comparison given information on eating disorders was only collected in one of the three survey waves and only a small proportion (4.2%) reported having received a diagnosis (**Table S1**). There were no notable overall differences by ethnicity, socioeconomic markers, smoking status, or level of alcohol consumption. However, among men, prevalence appeared to be higher among those from less advantaged socioeconomic groups (i.e., those from occupational social grades C2DE, those finding it difficult to manage financially, and those not in work due to long-term illness or disability), whereas prevalence was more similar or showed the opposite pattern among women (**Table S2**).

**Table 2.** Use of GLP-1 receptor agonists to support weight loss, and interest in using weight-loss medication, by participant characteristics.

|  | Used GLP-1RAs to support weight loss in the past year<sup>1</sup> |  | Interested in using weight-loss medication in the next year<sup>2</sup> |  |
| --- | --- | --- | --- | --- |
|  | % [95% CI] | OR [95% CI]<sup>3</sup> | % [95% CI] | OR [95% CI]<sup>3</sup> |
| Gender |  |  |  |  |
| Man | 1.7 [1.1–2.3] | Ref | 5.1 [4.2–6.1] | Ref |
| Woman | 4.0 [3.2–4.8] | 2.40 [1.62–3.56] | 8.9 [7.7–10.1] | 1.85 [1.44–2.38] |
| Age (years)<sup>4</sup> |  |  |  |  |
| 18 | 1.2 [0.7–2.1] | Ref | 5.5 [3.8–7.9] | Ref |
| 25 | 1.8 [1.2–2.7] | 1.53 [1.31–1.74] | 6.8 [5.3–8.6] | 1.26 [1.11–1.40] |
| 35 | 3.1 [2.4–3.9] | 2.64 [2.03–3.26] | 8.7 [7.5–10.0] | 1.67 [1.30–2.04] |
| 45 | 4.2 [3.4–5.3] | 3.74 [2.64–4.85] | 9.7 [8.4–11.3] | 1.91 [1.35–2.46] |
| 55 | 4.2 [3.4–5.2] | 3.71 [2.42–4.99] | 8.6 [7.3–10.1] | 1.66 [1.07–2.25] |
| 65 | 2.9 [2.2–3.7] | 2.46 [1.46–3.46] | 5.8 [4.8–6.9] | 1.07 [0.58–1.56] |
| 75 | 1.5 [1–2.4] | 1.27 [0.51–2.03] | 3.2 [2.4–4.4] | 0.57 [0.12–1.02] |
| Ethnicity |  |  |  |  |
| White | 2.9 [2.4–3.4] | Ref | 6.6 [5.8–7.4] | Ref |
| Minority ethnic group | 2.9 [1.5–4.2] | 1.05 [0.64–1.73] | 9.6 [7.0–12.3] | 1.30 [0.91–1.86] |
| Occupational social grade |  |  |  |  |
| ABC1 (more advantaged) | 3.1 [2.5–3.7] | Ref | 6.8 [5.9–7.8] | Ref |
| C2DE (less advantaged) | 2.6 [1.7–3.4] | 0.85 [0.57–1.27] | 7.4 [6.0–8.7] | 1.09 [0.84–1.41] |
| Financial situation |  |  |  |  |
| Living comfortably | 2.9 [2.1–3.7] | Ref | 4.5 [3.4–5.7] | Ref |
| Doing alright | 2.6 [1.8–3.4] | 0.88 [0.57–1.34] | 6.4 [5.1–7.7] | 1.26 [0.89–1.78] |
| Just about getting by | 2.9 [1.8–4.1] | 0.99 [0.61–1.62] | 8.7 [7.0–10.5] | 1.98 [1.40–2.80] |
| Finding it quite difficult | 3.3 [1.5–5.1] | 1.11 [0.58–2.12] | 9.6 [6.6–12.6] | 2.02 [1.28–3.19] |
| Finding it very difficult | 3.7 [1.2–6.3] | 1.17 [0.55–2.50] | 11.7 [7.6–15.7] | 2.01 [1.16–3.48] |
| Economically inactive due to long-term illness or disability |  |  |  |  |
| No | 2.8 [2.3–3.3] | Ref | 6.7 [5.9–7.5] | Ref |
| Yes | 4.3 [1.7–6.9] | 1.26 [0.66–2.43] | 13.4 [9.1–17.7] | 1.85 [1.19–2.88] |
| Smoking status |  |  |  |  |
| Never | 2.6 [2.0–3.3] | Ref | 6.4 [5.4–7.4] | Ref |
| Former | 3.6 [2.6–4.5] | 1.35 [0.92–1.97] | 8.1 [6.5–9.6] | 1.26 [0.93–1.71] |
| Current | 2.4 [1.2–3.7] | 0.87 [0.48–1.56] | 7.5 [5.3–9.6] | 1.09 [0.76–1.56] |

**Table 2. continued**
|  | Used GLP-1RAs to support weight loss in the past year<sup>1</sup> |  | Interested in using weight-loss medication in the next year<sup>2</sup> |  |
| --- | --- | --- | --- | --- |
|  | % [95% CI] | OR [95% CI]<sup>3</sup> | % [95% CI] | OR [95% CI]<sup>3</sup> |
| Level of alcohol consumption (AUDIT-C)<sup>5</sup> |  |  |  |  |
| 0 | 3.2 [2.4–4.3] | Ref | 8.3 [7.0–9.9] | Ref |
| 3 | 2.5 [2.0–3.3] | 0.79 [0.41–1.17] | 6.0 [5.0–7.2] | 0.76 [0.49–1.02] |
| 5 | 2.5 [1.9–3.3] | 0.82 [0.39–1.26] | 5.9 [4.9–7.1] | 0.77 [0.46–1.07] |
| 8 | 3.0 [2.2–4.1] | 1.12 [0.66–1.58] | 7.4 [6.0–9.0] | 0.99 [0.64–1.33] |
| 11 | 3.9 [2.0–7.4] | 1.67 [0.92–2.42] | 10.0 [6.6–14.8] | 1.40 [0.81–1.99] |
| Past-month psychological distress |  |  |  |  |
| None/low | 2.4 [1.8–2.9] | Ref | 5.2 [4.3–6.1] | Ref |
| Moderate/severe | 3.7 [2.8–4.6] | 1.62 [1.13–2.32] | 10.0 [8.5–11.4] | 1.81 [1.40–2.35] |
| History of eating disorders<sup>6</sup> |  |  |  |  |
| No | 2.1 [1.4–2.9] | Ref | 7.3 [5.8–8.7] | Ref |
| Yes | 4.4 [0.1–8.8] | 2.12 [0.69–6.53] | 13.2 [4.0–22.4] | 1.59 [0.66–3.82] |

CI, confidence interval. OR, odds ratio.
<sup>1</sup> Among adults in Great Britain.
<sup>2</sup> Among those who had not used a GLP-1RA or other medication for weight loss in the past year.
<sup>3</sup> Adjusted for age and gender (analyses by age are adjusted for gender only and analyses by gender are adjusted for age only).
<sup>4</sup> Predicted estimates from logistic regression models with age modelled using restricted cubic splines. Note that the models used to derive these estimates included data from participants of all ages, not only those who were aged exactly 16, 25, 35, 45, 55, or 65 years.
<sup>5</sup> Predicted estimates from logistic regression models with AUDIT-C score modelled using restricted cubic splines. Note that the models used to derive these estimates included data from all participants who provided data on AUDIT-C, not only those who scored exactly 0, 3, 5, 8, or 11.
<sup>6</sup> History of eating disorders was only assessed in February 2025; analyses were restricted to participants surveyed in this wave.
Estimates stratified by gender are provided in **Table S2** (use in the past year) and **Table S4** (interest).

### Types of GLP-1RAs being used

The most commonly used GLP-1RA was Mounjaro (tirzepatide; 2.3%), followed by Ozempic (semaglutide; 1.3%), Wegovy (semaglutide; 0.8%), Rybelsus (semaglutide; 0.6%), and Saxenda (liraglutide; 0.4%; **Table 1**). Of note, Mounjaro was most commonly used to support weight loss; it was three times more prevalent for this reason than the next most popular GLP-1RA (2.0% vs. Wegovy at 0.6%; **Table 1**).

Among those who used a GLP-1RA for weight loss in the past year, 85.0% [79.3–90.7%] used a medication licensed for weight loss in Great Britain – 69.5% [62.0–67.0%] reported using Mounjaro, 20.0% [12.9–27.0%] Wegovy, and 3.3% [0.0–6.8%] Saxenda (note that participants could select multiple medications, so these values sum to more than the total; 19.4% [12.7–26.1%] of those who reported using GLP-1RAs for any reason reported using more than one GLP-1RA – 0.7% [0.5– 1.0%] of all participants). Among those who used a GLP-1RA exclusively for weight loss, those numbers were 91.4% [85.6–97.2%], 80.2% [71.9–88.6%], 21.4% [12.4–30.3%], and 0.9% [0.0–2.6%], respectively.

### Interest in using weight-loss medications

Among participants who had not used a GLP-1RA or other medication for weight loss in the past year (96.0% [95.5–96.6%] of the sample), 6.5% [5.7–7.3%] said they would be likely to consider using a weight-loss medication in the next year (2.5% [2.1–3.0%] very likely and 4.0% [3.4–4.6%] fairly likely; **Figure 1**). This is approximately 3.3 million adults in Great Britain (53.7 million adults x 96.0% not used medication to support weight loss in the past year x 6.5%), of whom around 1.3 million (53.7 million x 96.0% x 2.5%) are very likely to consider it.

**Figure 1.**
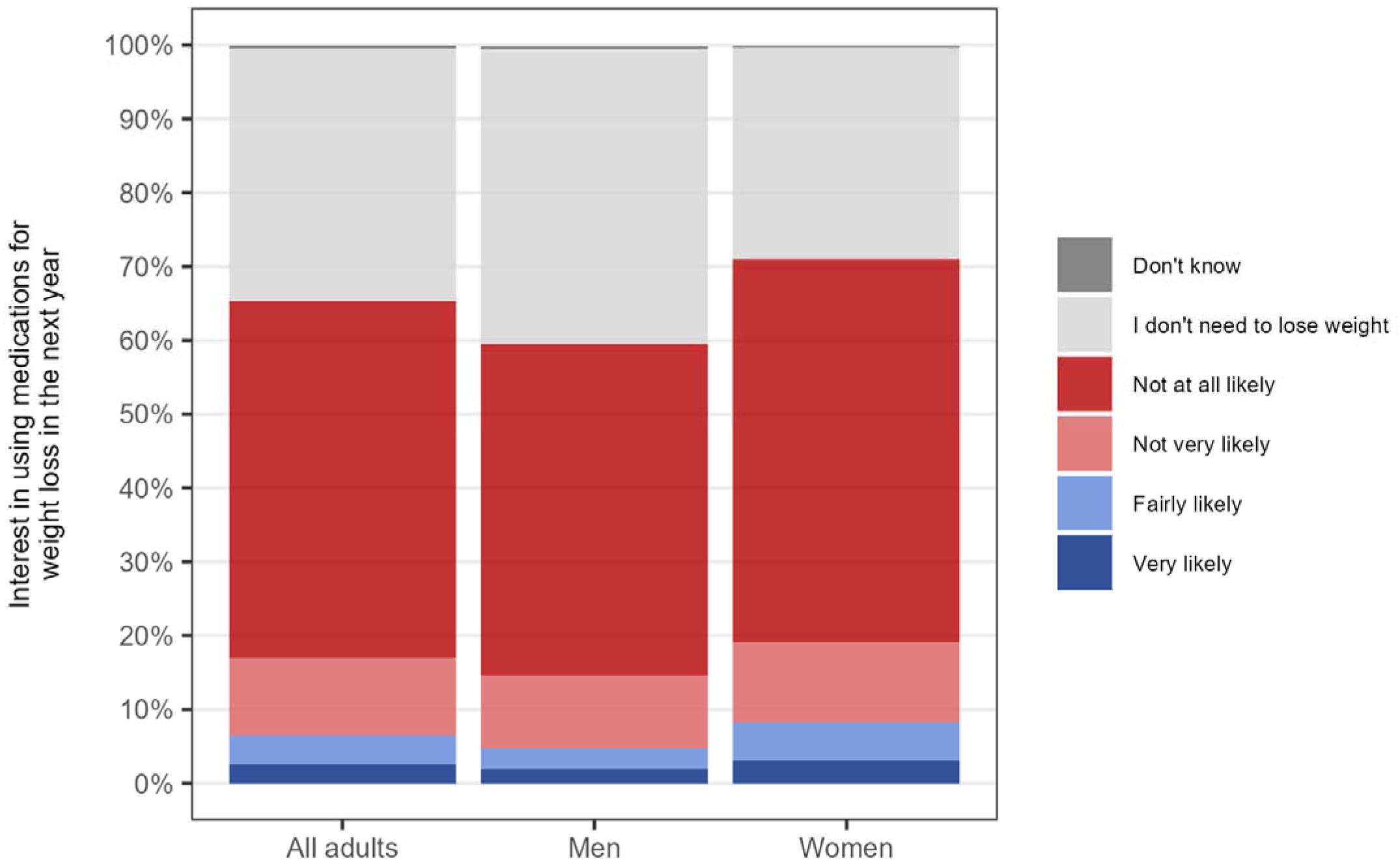
Interest in using weight-loss medication among adults (≥18y) in Great Britain who have not done so in the past year, overall and by gender

Interest in using weight-loss medications differed according to participant characteristics (**Table 2**). Some of these differences mirrored patterns observed for past-year use of GLP-1RAs for weight loss. The proportion who said they would be likely to consider using weight-loss medications in the next year was higher among women than men (8.9% vs. 5.1%; OR=1.85 [1.44–2.38]; **Figure 1**), those in mid-life (e.g., 9.7% among those aged 45 vs. 5.5% and 3.2% among those aged 18 and 75, respectively), and those who reported moderate/severe psychological distress (10.0% vs. 5.2% among those reporting no/low distress; OR=1.81 [1.40–2.35]). It also appeared to be higher among those who reported a history of eating disorders (13.2% vs. 7.3% among those who did not; OR=1.59 [0.66-3.82]), but again, there was substantial imprecision.

There were also some subgroup differences that were not observed for past-year use (**Table 2**). Interest in using weight-loss medications in the next year was higher among those in less favourable financial situations (e.g., 11.7% among those who reported finding it very difficult to manage financially vs. 4.5% among those living comfortably; OR=2.01 [1.16–3.48]) and among those who were not in work due to long-term illness or disability (13.4% vs. 6.7% of those not in this situation; OR=1.85 [1.19–2.88]). It also appeared to be slightly higher among those from minority ethnic groups (9.6% vs. 6.6% among white participants; OR=1.30 [0.91–1.86]), but this difference was uncertain. There were no notable differences by occupational social grade, smoking status, or level of alcohol consumption.

## Discussion

Our data suggest that in the first quarter of 2025, 4.9 million adults in Great Britain – nearly one in ten – either had recently used medication to support weight loss or were interested in doing so in the near future. Of these, approximately 1.6 million adults used GLP-1RAs for weight management, with 910,000 (∼60%) using them exclusively for this purpose. The majority reported using GLP-1RAs that are licensed for weight loss in Great Britain, most commonly Mounjaro (tirzepatide). Use and interest were more prevalent among women, people in mid-life, and those reporting past-month psychological distress, and also appeared higher among those with a history of eating disorders. Interest was also higher among people facing greater socioeconomic disadvantage, including those in financial difficulty or unable to work due to long-term illness or disability.

These data provide important insights into the emerging landscape of GLP-1RA use and potential future demand in Great Britain. The substantial level of current use, combined with even greater levels of interest, highlights growing public awareness of pharmacological options for weight management. However, the gap between interest and current use suggests there may be unmet demand. However, in the absence of data on people’s underlying medical risk factors, it is not possible to understand the extent to which this reflects ‘true’ medical need, e.g. meeting either current NICE criteria for weight loss medication or a lower threshold at which health benefits outweigh the harms. While the numbers we estimate are for Great Britain as a whole, it is noteworthy that the number using these medications for weight loss far exceeds the initial expectation for NHS England to provide treatment for 220,000 people in the first three years, which is likely to have important consequences for NHS budgets, although our estimate is less than the number eligible according to NICE guidelines (3.4 million).^14^ Part of the discrepancy may reflect prescribing outside the NHS. It is unclear to what extent this use may be driven by health concerns – and falls within either NICE recommendations or licensed use – or desire to lose weight for other reasons. Nonetheless, this trend is consistent with broader societal shifts towards medicalised approaches to managing obesity^11,40^ and points to a need for healthcare systems and policymakers to decide how best to manage a continued surge in demand.

Consistent with previous studies showing greater uptake of weight-loss treatments among women,^24^ we observed higher use and interest among women than men. This may reflect a combination of factors, including greater social pressures on women regarding body image and weight,^25^ as well as gender differences in healthcare-seeking behaviour.^41^ Women have much higher prevalence of severe obesity than men,^2^ and are typically more likely to engage with health (and preventive) services,^42^ engage with weight management services and discuss weight loss with healthcare professionals,^24^ which may facilitate greater access to emerging treatments like GLP-1RAs.

We also identified socioeconomic patterns. Although current use of GLP-1RAs for weight loss was relatively consistent across socioeconomic groups, this did vary by gender. Among men, use was more common in less advantaged groups, while among women, use was relatively consistent across socioeconomic strata. This may reflect higher prevalence of obesity-related metabolic comorbidities among men compared to women, which may also be socially patterned.^43,44^ For men, from a health inequalities perspective it is encouraging that uptake is higher among people from less advantaged groups. It is also unusual for the adoption of an expensive new medication or new medical technology, and this finding warrants further scrutiny and exploration. For women, one possible explanation is that women may be more likely than men to access GLP-1RAs through private routes, which often require substantial out-of-pocket costs^19^ and may be less accessible to those with fewer financial resources. Further research is needed to explore these findings more fully.

Interest in future use was higher among those experiencing financial hardship or unemployment due to long-term illness or disability. The greater interest among groups disproportionately affected by severe obesity^2,45^ and its related comorbidities may reflect a recognition of the potential health benefits of weight-loss medications for these people.^27,28^ It may also reflect a lack of perceived viable alternatives to lose weight: lifestyle interventions for weight management (even very intensive, well-designed, multicomponent ones) have a very modest impact on weight loss.^46,47^

Use of GLP-1RAs for weight loss and interest in future use were also higher among those experiencing psychological distress and those reporting a history of eating disorders. This may reflect the well-documented bidirectional relationship between mental health conditions – such as depression, anxiety, and binge eating disorder – and obesity.^48,49^ Mental health disorders are also more prevalent among women,^50^ which may contribute to observed gender differences in GLP-1RA use and interest. Individuals with higher levels of psychological distress may be more vulnerable to weight-related stigma or internalised weight bias,^51^ leading to greater concern about appearance and heightened motivation to seek pharmacological interventions. Greater body shame, weight concerns, anti-fat bias, and disordered eating behaviours have all been linked to greater interest in using GLP-1RAs.^52^ However, the direction of these associations is unclear and may be bidirectional. Emerging evidence has raised concerns about potential adverse mental health effects associated with GLP-1RA use, including increased psychological distress or suicidal ideation in some users, although data are currently limited and inconclusive.^53,54^ These patterns highlight the importance of further research to better understand the relationship between mental health and both actual and intended use of GLP- 1RAs for weight management.

Another notable finding is the substantial proportion of past-year users who reported using GLP- 1RAs that are not licensed for weight loss (15.0% of all those who reported using a GLP-1RA to support weight loss; 8.6% of those reporting use exclusively for weight loss), indicating off-label prescribing^55^ or procurement through non-medical channels.^56,57^ This may highlight potential issues in prescribing practices, patient understanding, and/or regulatory oversight. Some off-label prescribing should be expected: in the UK, liraglutide has an indication for weight loss only, but NICE guidelines list it as a treatment option for type 2 diabetes.^58^ While off-license prescribing can be common in some areas of medicine,^59^ it can also pose safety risks^60^ – particularly when medications are accessed without appropriate clinical supervision. There are concerns about GLP1-RA being purchased without a prescription through unregulated or illicit channels, with very light medical supervision or being used outside their licensed use (i.e. at lower BMI). Further work is needed, but this data raises further questions about their actual use and whether appropriate safety standards are in place and being adhered to. Depending on how these medications are being accessed, public health messaging warning of the potential harms of purchasing online or from other non-medical outlets may also be helpful.

The rising demand for GLP-1RAs to support weight loss presents significant challenges for healthcare systems such as the NHS. Responding to this demand safely and equitably, while focusing on those with medical need, will require difficult decisions about access, prioritisation, and resource allocation. These discussions are already underway and have led to a new proposition about how to identify those with clinical need who would benefit most from these drugs, although this is not without controversy.^61,62^ Although widespread adoption of GLP-1RAs has the potential to improve population health by reducing obesity-related diseases, the financial implications are considerable. These medications are expensive,^11^ are likely to require long-term use, and scaling up provision could place a heavy burden on NHS budgets already strained by the management of chronic conditions and recent structural reorganisations.^63^

Equally the majority of the population thought they were not interested in using the medications, and while attitudes may change, as with other medical interventions this might suggest limits on public willingness to use them. While GLP-1RAs are generally considered cost-effective for treating obesity in targeted populations,^64–66^ alternative strategies – such as expanding bariatric surgery access, enhancing lifestyle intervention programmes, pharmaceutical management of cardiometabolic comorbidities, or population-level policies should also be considered. These may not yet have achieved an equivalent scale of impact compared to GLP-1RAs, but some have not been tried at scale or with the intensity that is required, but may offer more cost-effective, long lasting or equitable solutions.^11^ Policymakers will need to balance the benefits of increasing GLP-1RA access against broader public health investments, which have been slow to be implemented (e.g., advertising and product promotion restrictions^63^ that are not yet in place, and which have been further delayed). Furthermore, it will be essential to ensure that access is equitable, clinically justified, and aligned with evidence-based guidelines to prevent widening health inequalities or encouraging inappropriate prescribing. In addition, there is a need to ensure these medications remain available to those who need them for reasons other than weight loss (over a third of users in 2024, according to our data; ∼1.5 million people).

A key strength of this study is the relatively large, nationally representative sample. In addition, the inclusion of demographic, socioeconomic, and psychological factors provides a detailed picture of patterns of use and potential demand within different population subgroups. There were also several limitations. All data were self-reported and relied on recall of the past year, introducing scope for bias. Given the cross-sectional nature of the study, we were unable to disentangle directional associations with time-varying sample characteristics and may have been unable to detect some such associations. For example, while we did not observe an association between alcohol consumption and past-year use of GLP-1RAs, it is possible that those who used these medications initially had higher consumption but experienced reductions in alcohol consumption between initiating medication use and completing the survey.^8^ Due to limited availability of funding for the GLP-1RA survey items, we were unable to collect more detailed information about how and why people were accessing these medications. No data were available on height and weight, so we were unable to explore differences by BMI status, and cannot make assessments about the appropriateness of use against medical criteria for use. We also did not ask specifically about all GLP-1RAs licensed in the UK, which may have resulted in incomplete information on the types of medications used and caused us to underestimate the overall prevalence of use. Further research is needed to explore the source of GLP-1RAs (e.g. NHS, private prescription in person, post/internet with a prescription, post/internet with no formal prescription) and to assess the appropriateness of use (i.e., the extent to which people who are using these drugs for weight loss fall within the indications of the NICE guidelines). Regularly including these types of questions in a representative, repeat cross-sectional household survey like the Smoking Toolkit Study would offer insights into how population-level use of, and demand for, GLP-1RA medications for weight loss is evolving over time. Qualitative research is also important for a richer understanding of people’s experiences of accessing and using these medications.

In conclusion, this study highlights substantial demand for GLP-1RAs to support weight loss in Great Britain. As drugs become more available, equitable access to these treatments should be prioritised to meet the needs of all population groups, particularly those who may face barriers to access despite high interest. At the same time, healthcare systems must be prepared for the potential pressures on capacity and budgets. To support safe, effective, and equitable use, there is a clear need for regular, population-level monitoring – not only of the prevalence and patterns of use, but also of access, appropriateness, health outcomes, and broader system impacts. Such surveillance will be essential to inform responsive healthcare planning, guide and evaluate policy decisions, and ensure that these treatments deliver sustainable benefits without widening health inequalities or overburdening healthcare resources.

## Data Availability

Data produced in the present study are available upon reasonable request to the authors

## Declarations

### Ethics approval

Ethical approval for the STS was granted originally by the UCL Ethics Committee (ID 0498/001). Participants provide informed consent to take part in the study, and all methods are carried out in accordance with relevant regulations. The data are not collected by UCL and are anonymised when received by UCL.

## Competing interests

LS has acted as paid reviewer for grant awarding bodies and as a paid consultant for health care companies.

## Funding

This work was supported by Cancer Research UK (PRCRPG-Nov21\100002). JB is a member of the Behavioural Research UK Leadership Hub which is supported by the Economic and Social Research Council (ES/Y001044/1). For the purpose of Open Access, the author has applied a CC BY public copyright licence to any Author Accepted Manuscript version arising from this submission.

## References

1. NHS Digital. Statistics on Obesity, Physical Activity and Diet, England 2021. 2021 https://digital.nhs.uk/data-and-information/publications/statistical/statistics-on-obesity-physical-activity-and-diet/england-2021/part-1-obesity-related-hospital-admissions (accessed 10 Mar2025).

2. NHS Digital. Health Survey for England, 2022 Part 2. 2024 https://digital.nhs.uk/data-and-information/publications/statistical/health-survey-for-england/2022-part-2/adult-overweight-and-obesity (accessed 10 Mar2025).

3 Pan X-H, Tan B, Chin YH, Lee ECZ, Kong G, Chong B et al. Efficacy and safety of tirzepatide, GLP-1 receptor agonists, and other weight loss drugs in overweight and obesity: a network meta-analysis. Obesity 2024; 32: 840–856.

4 Wong HJ, Sim B, Teo YH, Teo YN, Chan MY, Yeo LLL et al. Efficacy of GLP-1 Receptor Agonists on Weight Loss, BMI, and Waist Circumference for Patients With Obesity or Overweight: A Systematic Review, Meta-analysis, and Meta-regression of 47 Randomized Controlled Trials. Diabetes Care 2025; **48**: 292–300.

5 Guo H, Yang J, Huang J, Xu L, Lv Y, Wang Y et al. Comparative efficacy and safety of GLP-1 receptor agonists for weight reduction: A model-based meta-analysis of placebo-controlled trials. Obes Pillars 2025; 13: 100162.

6 Lincoff AM, Brown-Frandsen K, Colhoun HM, Deanfield J, Emerson SS, Esbjerg S et al. Semaglutide and Cardiovascular Outcomes in Obesity without Diabetes. N Engl J Med 2023; 389: 2221–2232.

7 Marso SP, Bain SC, Consoli A, Eliaschewitz FG, Jódar E, Leiter LA et al. Semaglutide and Cardiovascular Outcomes in Patients with Type 2 Diabetes. N Engl J Med 2016; 375: 1834– 1844.

8 Martinelli S, Mazzotta A, Longaroni M, Petrucciani N. Potential role of glucagon-like peptide-1 (GLP-1) receptor agonists in substance use disorder: A systematic review of randomized trials. Drug Alcohol Depend 2024; 264: 112424.

9 Thomsen RW, Mailhac A, Løhde JB, Pottegård A. Real-world evidence on the utilization, clinical and comparative effectiveness, and adverse effects of newer GLP-1RA-based weight-loss therapies. Diabetes Obes Metab 2025; 27: 66–88.

10. Sridhar D. We know so little about taking weight-loss drugs without prescription – is it really worth it? The Guardian. 2025. https://www.theguardian.com/commentisfree/2025/feb/25/weight-loss-drugs-without-prescription (accessed 4 Mar2025).

11 Mytton OT, Head V, Reckless I. Public health perspective on new weight loss medications. BMJ 2024; 384: q196.

12. Medicines and Healthcare products Regulatory Agency. GLP-1 receptor agonists: reminder of the potential side effects and to be aware of the potential for misuse. GOV.UK. 24 Oct 24.https://www.gov.uk/drug-safety-update/glp-1-receptor-agonists-reminder-of-the-potential-side-effects-and-to-be-aware-of-the-potential-for-misuse (accessed 4 Mar2025).

13. National Institute for Health and Care Excellence. Overweight and obesity management | NICE guideline | NG246. 2025. https://www.nice.org.uk/guidance/ng246/chapter/Medicines-and-surgery (accessed 21 May2025).

14. NHS England. Interim commissioning guidance: Implementation of the NICE Technology Appraisal TA1026 and the NICE funding variation for tirzepatide (Mounjaro®) for the management of obesity. 2025 https://www.england.nhs.uk/publication/interim-commissioning-guidance-implementation-of-the-nice-technology-appraisal-ta1026-and-the-nice-funding-variation-for-tirzepatide-mounjaro-for-the-management-of-obesity/ (accessed 15 May2025).

15. Poll: 1 in 8 Adults Say They’ve Taken a GLP-1 Drug, Including 4 in 10 of Those with Diabetes and 1 in 4 of Those with Heart Disease. KFF. 2024. https://www.kff.org/health-costs/press-release/poll-1-in-8-adults-say-theyve-taken-a-glp-1-drug-including-4-in-10-of-those-with-diabetes-and-1-in-4-of-those-with-heart-disease/ (accessed 4 Mar2025).

16. Correspondent PK Health. NHS struggling to cope with ‘overwhelming’ weight-loss drug demand. 2024. https://www.thetimes.com/uk/healthcare/article/nhs-weight-loss-drugs-mounjaro-ozempic-jq9n825pn (accessed 4 Mar2025).

17. Semaglutide (British National Formulary code 0601023AW). OpenPrescribing. https://openprescribing.net/chemical/0601023AW/ (accessed 27 Mar2025).

18. Tirzepatide (British National Formulary code 0601023AZ). OpenPrescribing. https://openprescribing.net/chemical/0601023AZ/ (accessed 27 Mar2025).

19. Campbell D, editor DCH policy. One in five Britons would use weight-loss drug if free on NHS, poll reveals. The Guardian. 2024. https://www.theguardian.com/society/2024/dec/28/one-in-five-britons-weight-loss-drug-free-nhs-poll (accessed 4 Mar2025).

20 Sheth K, Garza E, Saju A, Nazir N, Agarwal A. Wernicke Encephalopathy Associated With Semaglutide Use. Cureus; 16: e61783.

21 Ali SA, Khadra M, Sitto M, Graifman M, Gietzen J. S4686 Semaglutide’s Hidden Perils: A Rare Case of Malnutrition and Wernicke Encephalopathy. Off J Am Coll Gastroenterol ACG 2024; **119**: S2964.

22 Sharma N, Vura NVRK, Shweikeh F, Ramirez-Osoria LC, Chakinala RC, Sharma AN. S4690 Semaglutide-Linked Dry Beriberi: A Rare Adverse Reaction. Off J Am Coll Gastroenterol ACG 2024; **119**: S2967.

23. Foster S, Kyle A. From Weight Loss to Neurological Deficits: A Case of Wernicke’s Encephalopathy Stemming From Prescription Weight Loss Medication. EM Resid. 2023. https://www.emra.org/emresident/article/wernicke-july-2023 (accessed 27 Mar2025).

24 Cooper AJ, Gupta SR, Moustafa AF, Chao AM. Sex/Gender Differences in Obesity Prevalence, Comorbidities, and Treatment. Curr Obes Rep 2021; 10: 458–466.

25 Murnen SK, Don BP. Body image and gender roles. In: Encyclopedia of body image and human appearance. Academic Press: San Diego, 2012, pp 128–134.

26 Dellgren JL, Persad G, Emanuel EJ. International coverage of GLP-1 receptor agonists: a review and ethical analysis of discordant approaches. The Lancet 2024; 404: 902–906.

27. Stiebahl S. Obesity statistics. 2025. https://commonslibrary.parliament.uk/research-briefings/sn03336/ (accessed 10 Mar2025).

28 El-Sayed AM, Scarborough P, Galea S. Unevenly distributed: a systematic review of the health literature about socioeconomic inequalities in adult obesity in the United Kingdom. BMC Public Health 2012; 12: 18.

29 Fidler JA, Shahab L, West O, Jarvis MJ, McEwen A, Stapleton JA et al. ‘The smoking toolkit study’: a national study of smoking and smoking cessation in England. BMC Public Health 2011; 11: 479.

30 Kock L, Shahab L, Moore G, Beard E, Bauld L, Reid G, et al. Protocol for expansion of an existing national monthly survey of smoking behaviour and alcohol use in England to Scotland and Wales: The Smoking and Alcohol Toolkit Study. Wellcome Open Res 2021; 6: 67.

31 Jackson SE, Tattan-Birch H, Shahab L, Brown J. Trends in long term vaping among adults in England, 2013-23: population based study. BMJ 2024; 386: e079016.

32. National Readership Survey. Social grade - definitions and discriminatory power. 2007. http://www.nrs.co.uk/lifestyle.html (accessed 1 Oct2012).

33 Downward P, Rasciute S, Kumar H. Health, subjective financial situation and well-being: a longitudinal observational study. Health Qual Life Outcomes 2020; 18: 203.

34 Rumpf H-J, Hapke U, Meyer C, John U. Screening for alcohol use disorders and at-risk drinking in the general population: psychometric performance of three questionnaires. Alcohol Alcohol 2002; 37: 261–268.

35 Kessler RC, Andrews G, Colpe LJ, Hiripi E, Mroczek DK, Normand S-LT et al. Short screening scales to monitor population prevalences and trends in non-specific psychological distress. Psychol Med 2002; 32: 959–976.

36 Kessler RC, Green JG, Gruber MJ, Sampson NA, Bromet E, Cuitan M et al. Screening for serious mental illness in the general population with the K6 screening scale: results from the WHO World Mental Health (WMH) survey initiative. Int J Methods Psychiatr Res 2010; 19: 4–22.

37 Prochaska JJ, Sung H-Y, Max W, Shi Y, Ong M. Validity study of the K6 scale as a measure of moderate mental distress based on mental health treatment need and utilization. Int J Methods Psychiatr Res 2012; 21: 88–97.

38. Rubin DB. Multiple imputation. In: Flexible Imputation of Missing Data, Second Edition. Chapman and Hall/CRC, 2018.

39. Office for National Statistics. Population estimates for the UK, England, Wales, Scotland, and Northern Ireland: mid-2022. 2024. https://www.ons.gov.uk/peoplepopulationandcommunity/populationandmigration/populationestimates/bulletins/annualmidyearpopulationestimates/mid2022 (accessed 2 May2024).

40 Rose G. Sick Individuals and Sick Populations. Int J Epidemiol 1985; 14: 32–38.

41 Wang Y, Hunt K, Nazareth I, Freemantle N, Petersen I. Do men consult less than women? An analysis of routinely collected UK general practice data. BMJ Open 2013; 3: e003320.

42 Bertakis KD, Azari R, Helms LJ, Callahan EJ, Robbins JA. Gender Differences in the Utilization of Health Care Services. J Fam Pract 2000; 49: 147–152.

43 Koceva A, Herman R, Janez A, Rakusa M, Jensterle M. Sex- and Gender-Related Differences in Obesity: From Pathophysiological Mechanisms to Clinical Implications. Int J Mol Sci 2024; 25: 7342.

44. The Health Foundation. Inequalities in life expectancy and healthy life expectancy. 2025. https://www.health.org.uk/evidence-hub/health-inequalities/inequalities-in-life-expectancy-and-healthy-life-expectancy (accessed 27 May2025).

45 Booth HP, Charlton J, Gulliford MC. Socioeconomic inequality in morbid obesity with body mass index more than 40 kg/m2 in the United States and England. SSM - Popul Health 2017; 3: 172–178.

46 Singh N, Stewart RAH, Benatar JR. Intensity and duration of lifestyle interventions for long-term weight loss and association with mortality: a meta-analysis of randomised trials. BMJ Open 2019; 9: e029966.

47 Madigan CD, Graham HE, Sturgiss E, Kettle VE, Gokal K, Biddle G et al. Effectiveness of weight management interventions for adults delivered in primary care: systematic review and meta-analysis of randomised controlled trials. BMJ 2022; 377: e069719.

48 Avila C, Holloway AC, Hahn MK, Morrison KM, Restivo M, Anglin R et al. An Overview of Links Between Obesity and Mental Health. Curr Obes Rep 2015; 4: 303–310.

49 Milaneschi Y, Simmons WK, van Rossum EFC, Penninx BW. Depression and obesity: evidence of shared biological mechanisms. Mol Psychiatry 2019; 24: 18–33.

50 Piccinelli M, Wilkinson G. Gender differences in depression. Critical review. Br J Psychiatry J Ment Sci 2000; 177: 486–492.

51 Pudney EV, Himmelstein MS, Puhl RM, Foster GD. Distressed or not distressed? A mixed methods examination of reactions to weight stigma and implications for emotional wellbeing and internalized weight bias. Soc Sci Med 2020; 249: 112854.

52 Markey CH, August KJ, Malik D, Richeson A. Body image and interest in GLP-1 weight loss medications. Body Image 2025; 53: 101890.

53 McIntyre RS. Glucagon-like peptide-1 receptor agonists (GLP-1 RAs) and suicidality: what do we know and future vistas. Expert Opin Drug Saf 2024; 23: 539–542.

54 Salvo F, Faillie J-L. GLP-1 Receptor Agonists and Suicidality—Caution Is Needed. JAMA Netw Open 2024; 7: e2423335.

55 Aronson JK, Ferner RE. Unlicensed and off-label uses of medicines: definitions and clarification of terminology. Br J Clin Pharmacol 2017; 83: 2615–2625.

56 Basch CH, Yousaf H, Hillyer GC. Online purchasing options for GLP-1 agonists: Accessibility, marketing practices, and consumer safety concerns. J Med Surg Public Health 2025; 5: 100183.

57. Pearson SD, Whaley CM, Emond SK. Affordable Access to GLP-1 Obesity Medications: Strategies to Guide Market Action and Policy Solutions. Institute for Clinical and Economic Review, 2025 https://icer.org/wp-content/uploads/2025/04/Affordable-Access-to-GLP-1-Obesity-Medications-_-ICER-White-Paper-_-04.09.2025.pdf (accessed 28 Apr2025).

58. National Institute for Health and Care Excellence. Diabetes - type 2: GLP-1 receptor agonists. 2025. https://cks.nice.org.uk/topics/diabetes-type-2/prescribing-information/glp-1-receptor-agonists/ (accessed 27 May2025).

59 Eguale T, Buckeridge DL, Winslade NE, Benedetti A, Hanley JA, Tamblyn R. Drug, Patient, and Physician Characteristics Associated With Off-label Prescribing in Primary Care. Arch Intern Med 2012; 172: 781–788.

60 Van Norman GA. Off-Label Use vs Off-Label Marketing of Drugs. JACC Basic Transl Sci 2023; 8: 224–233.

61 Rubino F, Cummings DE, Eckel RH, Cohen RV, Wilding JPH, Brown WA et al. Definition and diagnostic criteria of clinical obesity. Lancet Diabetes Endocrinol 2025; 13: 221–262.

62 Mytton OT, Campbell-Scherer D, Reckless I, Llewellyn C. Diagnosing and defining obesity. BMJ 2025; 388: r460.

63. Department of Health and Social Care. Tackling obesity: empowering adults and children to live healthier lives. 2020 https://www.gov.uk/government/publications/tackling-obesity-government-strategy/tackling-obesity-empowering-adults-and-children-to-live-healthier-lives (accessed 23 Jun2021).

64 Evans M, Evans W, Godbeer F, Edgar L, Spaepen E, Davies AL. Analysis of Tirzepatide Acquisition Costs and Weight Reduction Outcomes in the United Kingdom: Insights from the SURMOUNT-1 Study. Adv Ther 2025. doi:10.1007/s12325-025-03194-8.

65 Kim N, Wang J, Burudpakdee C, Song Y, Ramasamy A, Xie Y et al. Cost-effectiveness analysis of semaglutide 2.4 mg for the treatment of adult patients with overweight and obesity in the United States. J Manag Care Spec Pharm 2022; 28: 740–752.

66 Hu Y, Zheng S-L, Ye X-L, Shi J-N, Zheng X-W, Pan H-S et al. Cost-effectiveness analysis of 4 GLP-1RAs in the treatment of obesity in a US setting. Ann Transl Med 2022; 10: 152.

