## Supplementary file for "Prevalence of use and interest in using glucagon-like peptide-1 receptor agonists for weight loss: a population study in Great Britain"

**Table S1.** Sample characteristics

| | Total eligible sample | Consented to participate in GLP-1RA module | Imputed (analysed) data[1] |
|---|---|---|---|
| Unweighted *n* | 5,893 | 5,260 | 5,893 |
| Age (years) | | | |
| Mean (SD) | 48.5 | 48.9 | 48.5 |
| 18-24 | 12.1 | 11.6 | 12.1 |
| 25-34 | 17.0 | 16.5 | 17.0 |
| 35-44 | 16.0 | 16.0 | 16.0 |
| 45-54 | 16.2 | 16.4 | 16.2 |
| 55-64 | 15.7 | 16.1 | 15.7 |
| ≥65 | 23.0 | 23.4 | 23.0 |
| *Missing* | 0 | 0 | 0 |
| Gender | | | |
| Man | 48.5 | 47.8 | 48.7 |
| Woman | 50.5 | 51.3 | 50.7 |
| In another way | 0.6 | 0.6 | 0.6 |
| *Missing* | 0.4 | 0.3 | 0 |
| Ethnicity | | | |
| White | 84.6 | 85.7 | 85.2 |
| Black | 4.8 | 4.6 | 4.9 |
| Asian | 5.2 | 4.8 | 5.3 |
| Mixed/multiple | 3.1 | 3.0 | 3.2 |
| Other | 1.5 | 1.3 | 1.5 |
| *Missing* | 0.7 | 0.6 | 0 |
| Occupational social grade | | | |
| ABC1 (more advantaged) | 55.9 | 56.3 | 55.9 |
| C2DE (less advantaged) | 44.1 | 43.7 | 44.1 |
| *Missing* | 0 | 0 | 0 |
| Financial situation | | | |
| Living comfortably | 27.0 | 27.4 | 27.2 |
| Doing alright | 32.5 | 32.5 | 32.7 |
| Just about getting by | 25.6 | 25.9 | 25.7 |
| Finding it quite difficult | 8.7 | 8.6 | 8.8 |
| Finding it very difficult | 5.6 | 5.2 | 5.6 |
| *Missing* | 0.6 | 0.4 | 0 |
| Economically inactive due to long-term illness or disability | | | |
| No | 94.0 | 94.0 | 94.4 |
| Yes | 5.6 | 5.5 | 5.6 |
| *Missing* | 0.5 | 0.4 | 0 |

*Table continues on next page.*

**Table S1.** *Continued*

| | Total eligible sample | Consented to participate in GLP-1RA module | Imputed (analysed) data[1] |
|---|---|---|---|
| Smoking status | | | |
| Never | 53.0 | 53.3 | 53.3 |
| Former | 29.9 | 30.7 | 30.0 |
| Current | 16.7 | 15.6 | 16.7 |
| *Missing* | 0.5 | 0.4 | 0 |
| Level of alcohol consumption (AUDIT-C) | | | |
| Mean (SD) | 3.2 | 3.3 | 3.2 |
| 0-4 | 67.4 | 67.3 | 69.0 |
| ≥5 | 30.3 | 30.6 | 31.0 |
| *Missing* | 2.4 | 2.1 | 0 |
| Past-month psychological distress | | | |
| None/low | 59.2 | 60.0 | 60.6 |
| Moderate/severe | 38.4 | 38.1 | 39.4 |
| *Missing* | 2.4 | 1.9 | 0 |
| History of eating disorders[2] | | | |
| No | 88.5 | 91.7 | 95.8 |
| Yes | 3.9 | 4.1 | 4.2 |
| *Missing* | 7.7 | 4.2 | 0 |

Data are presented as weighted percentages, unless otherwise specified.

[1] Missing data were imputed using multiple imputation with chained equations (5 imputed datasets).

[2] History of eating disorders was only assessed in February 2025 (*n*=1,961); missing cases do not include those surveyed in other waves.

### Gender-stratified results

#### *Prevalence of GLP-1RA use*

When stratified by gender (**Table S2**), use of GLP-1RAs to support weight loss was consistently higher among women than men across most subgroups. However, there were some differences in subgroup patterns. Among men, prevalence appeared to be higher among those from more disadvantaged socioeconomic groups (i.e., those from occupational social grades C2DE, those finding it difficult to manage financially, and those not in work due to long-term illness or disability), whereas prevalence was more similar or showed the opposite pattern among women. In addition, the association between eating disorders and higher prevalence of use of GLP-1RAs for weight loss appeared to be driven by men, with little difference by eating disorder history among women. However, we note that 95% CIs were wide, so these gender-stratified results should be interpreted with some caution. The types of GLP1-RAs used by men and women are summarised in **Table S3**.

#### *Interest in using weight-loss medications*

When stratified by gender (**Table S4**), interest in using weight-loss medications was consistently higher among women than men across most subgroups. However, there were some differences in subgroup patterns. Differences by age and ethnicity appeared more pronounced among women than men. Differences by financial difficulty, being out of work due to long-term illness or disability, and psychological distress appeared more pronounced among men. In addition, higher alcohol consumption was associated with greater interest in using weight-loss medications among women but not men. However, again, we advise some caution in interpreting these differences on account of the wide 95% CIs.

**Table S2.** Use of GLP-1 receptor agonists to support weight loss, by participant characteristics – stratified by gender

| | Used GLP-1RAs to support weight loss in the past year[1] | | | |
|---|---|---|---|---|
| | Men | | Women | |
| | % [95% CI] | OR [95% CI][2] | % [95% CI] | OR [95% CI][2] |
| Age (years)[3] | | | | |
| 18 | 0.6 [0.1–2.4] | Ref | 1.7 [0.9–3.3] | Ref |
| 25 | 0.9 [0.3–2.6] | 1.61 [1.12–2.10] | 2.6 [1.7–4.1] | 1.50 [1.27–1.74] |
| 35 | 1.7 [1.0–3.0] | 3.04 [1.44–4.64] | 4.3 [3.3–5.6] | 2.56 [1.89–3.23] |
| 45 | 2.5 [1.7–3.8] | 4.66 [1.54–7.78] | 5.9 [4.6–7.6] | 3.56 [2.42–4.70] |
| 55 | 2.6 [1.7–3.9] | 4.88 [1.20–8.56] | 5.8 [4.5–7.4] | 3.48 [2.25–4.71] |
| 65 | 1.8 [1.1–2.8] | 3.30 [0.62–5.99] | 3.9 [2.9–5.2] | 2.29 [1.38–3.20] |
| 75 | 0.9 [0.4–2.1] | 1.70 [0.00–3.43] | 2.0 [1.2–3.5] | 1.18 [0.40–1.95] |
| Ethnicity | | | | |
| White | 1.7 [1.1–2.3] | Ref | 4.0 [3.2–4.8] | Ref |
| Minority ethnic group | 1.6 [0.0–3.3] | 0.94 [0.32–2.81] | 4.0 [2.0–6.0] | 1.04 [0.58–1.85] |
| Occupational social grade | | | | |
| ABC1 (more advantaged) | 1.5 [0.8–2.2] | Ref | 4.6 [3.6–5.6] | Ref |
| C2DE (less advantaged) | 2.0 [0.9–3.1] | 1.30 [0.58–2.94] | 3.2 [2.0–4.5] | 0.73 [0.45–1.18] |
| Financial situation | | | | |
| Living comfortably | 1.8 [0.9–2.8] | Ref | 4.0 [2.6–5.4] | Ref |
| Doing alright | 1.3 [0.4–2.2] | 0.67 [0.28–1.62] | 3.7 [2.5–5.0] | 0.94 [0.55–1.58] |
| Just about getting by | 1.8 [0.4–3.2] | 0.95 [0.36–2.50] | 4.0 [2.4–5.6] | 0.99 [0.57–1.71] |
| Finding it quite difficult | 1.3 [0.0–3.2] | 0.63 [0.11–3.66] | 5.2 [2.2–8.1] | 1.24 [0.60–2.57] |
| Finding it very difficult | 3.9 [0.0–8.1] | 1.95 [0.61–6.27] | 3.7 [0.7–6.6] | 0.83 [0.33–2.13] |
| Economically inactive due to long-term illness or disability | | | | |
| No | 1.5 [1.0–2.1] | Ref | 4.0 [3.2–4.8] | Ref |
| Yes | 5.4 [0.8–10.0] | 3.02 [1.18–7.71] | 3.7 [0.7–6.7] | 0.71 [0.29–1.76] |
| Smoking status | | | | |
| Never | 1.5 [0.7–2.3] | Ref | 3.6 [2.7–4.6] | Ref |
| Former | 2.0 [1.0–3.1] | 1.36 [0.62–2.98] | 5.3 [3.7–6.8] | 1.44 [0.94–2.20] |
| Current | 1.8 [0.1–3.5] | 1.18 [0.40–3.51] | 3.0 [1.1–4.8] | 0.75 [0.37–1.54] |
| Level of alcohol consumption (AUDIT-C)[4] | | | | |
| 0 | 1.9 [1.0–3.5] | Ref | 4.1 [3.0–5.7] | Ref |
| 3 | 1.2 [0.7–2.1] | 0.63 [0.00–1.38] | 3.7 [2.8–5.0] | 0.89 [0.45–1.34] |
| 5 | 1.3 [0.7–2.3] | 0.65 [0.00–1.54] | 3.9 [2.9–5.1] | 0.90 [0.41–1.40] |
| 8 | 2.1 [1.4–3.3] | 1.06 [0.25–1.87] | 4.5 [2.8–7.2] | 1.02 [0.43–1.61] |
| 11 | 4.2 [1.8–9.4] | 2.05 [0.91–3.19] | 5.5 [2.0–13.9] | 1.19 [0.14–2.25] |

*Table continues on next page.*

**Table S2.** *continued*

| | Used GLP-1RAs to support weight loss in the past year[1] | | | |
|---|---|---|---|---|
| | Men | | Women | |
| | % [95% CI] | OR [95% CI][2] | % [95% CI] | OR [95% CI][2] |
| Past-month psychological distress | | | | |
| None/low | 1.5 [0.8–2.1] | Ref | 3.3 [2.4–4.2] | Ref |
| Moderate/severe | 2.2 [1.0–3.4] | 1.62 [0.77–3.42] | 4.9 [3.6–6.1] | 1.48 [0.98–2.24] |
| History of eating disorders[5] | | | | |
| No | 0.6 [0.1–1.1] | Ref | 3.7 [2.4–5.1] | Ref |
| Yes | 4.1 [0.0–12.3] | 9.96 [0.98–101.6] | 4.5 [0.0–9.6] | 1.14 [0.31–4.17] |

CI, confidence interval. OR, odds ratio.

[1] Among adults in Great Britain.

[2] Adjusted for age.

[3] Predicted estimates from logistic regression models with age modelled using restricted cubic splines. Note that the models used to derive these estimates included data from participants of all ages, not only those who were aged exactly 16, 25, 35, 45, 55, or 65 years.

[4] Predicted estimates from logistic regression models with AUDIT-C score modelled using restricted cubic splines. Note that the models used to derive these estimates included data from all participants who provided data on AUDIT-C, not only those who scored exactly 0, 3, 5, 8, or 11.

[5] History of eating disorders was only assessed in February 2025; analyses were restricted to participants surveyed in this wave.

**Table S3.** Past-year use of GLP-1 receptor agonists among men and women (≥18y) in Great Britain

| GLP-1RA used in past year | Authorised indication(s)[1] | Prevalence, % [95% CI] For any reason | To manage type 2 diabetes | To reduce the risk of heart disease | To support weight loss | Exclusively to support weight loss |
|---|---|---|---|---|---|---|
| **Men** | | | | | | |
| Any GLP-1RA listed below | - | 3.6 [2.7–4.5] | 1.7 [1.1–2.3] | 1.7 [1.2–2.3] | 1.7 [1.1–2.3] | 0.6 [0.3–1.0] |
| Saxenda, containing liraglutide | WL | 0.4 [0.0–0.7] | 0.2 [0.0–0.5] | 0.2 [0.0–0.4] | 0.1 [0.0–0.2] | - |
| Wegovy, containing semaglutide | WL, CRR | 0.6 [0.2–1.0] | 0.2 [0.0–0.4] | 0.3 [0.0–0.6] | 0.4 [0.1–0.8] | 0.2 [0.0–0.4] |
| Mounjaro, containing tirzepatide | WL, T2D | 1.2 [0.7–1.7] | 0.6 [0.3–0.9] | 0.5 [0.2–0.8] | 0.8 [0.4–1.2] | 0.4 [0.1–0.7] |
| Ozempic, containing semaglutide | T2D | 1.3 [0.7–1.8] | 0.6 [0.3–0.8] | 0.7 [0.3–1.1] | 0.4 [0.1–0.7] | 0.1 [0.0–0.2] |
| Rybelsus, containing semaglutide | T2D | 0.7 [0.3–1.1] | 0.6 [0.2–0.9] | 0.4 [0.1–0.7] | 0.4 [0.1–0.6] | 0.0 [0.0–0.1] |
| **Women** | | | | | | |
| Any GLP-1RA listed below | - | 5.3 [4.4–6.2] | 1.6 [1.1–2.2] | 1.6 [1.0–2.1] | 4.0 [3.2–4.8] | 2.8 [2.1–3.4] |
| Saxenda, containing liraglutide | WL | 0.3 [0.1–0.6] | 0.2 [0.0–0.4] | 0.1 [0.0–0.3] | 0.1 [0.0–0.3] | 0.0 [0.0–0.1] |
| Wegovy, containing semaglutide | WL, CRR | 0.9 [0.5–1.2] | 0.2 [0.0–0.5] | 0.3 [0.1–0.5] | 0.7 [0.4–1.0] | 0.5 [0.3–0.8] |
| Mounjaro, containing tirzepatide | WL, T2D | 3.3 [2.6–4.0] | 0.7 [0.4–1.1] | 0.8 [0.4–1.1] | 3.1 [2.4–3.8] | 2.3 [1.7–2.9] |
| Ozempic, containing semaglutide | T2D | 1.4 [0.8–2.1] | 0.7 [0.3–1.1] | 0.5 [0.1–0.9] | 0.7 [0.3–1.0] | 0.4 [0.1–0.6] |
| Rybelsus, containing semaglutide | T2D | 0.5 [0.1–0.8] | 0.4 [0.1–0.7] | 0.2 [0.0–0.4] | 0.3 [0.1–0.5] | 0.1 [0.0–0.2] |

CI, confidence interval. GLP-1RA, glucagon-like peptide-1 receptor agonist.
[1] Authorised indications for use in the UK. WL, weight loss. CRR, cardiovascular risk reduction. T2D, type 2 diabetes.
Data shown are weighted estimates of the proportion (with 95% CI) of men and women in Great Britain reporting past-year use of different GLP-1RAs, overall (i.e., for any reason) and stratified by the reason for use. Note that reasons are not mutually exclusive, except ‘exclusively to support weight loss’, which excludes participants reporting use for any other reason.
Empty cells indicate no participants reported use of the GLP-1RA for that reason.

**Table S4.** Interest in using weight-loss medication, by participant characteristics – stratified by gender

| | Used GLP-1RAs to support weight loss in the past year[1] | | | |
|---|---|---|---|---|
| | Men | | Women | |
| | % [95% CI] | OR [95% CI][2] | % [95% CI] | OR [95% CI][2] |
| Age (years)[3] | | | | |
| 18 | 4.4 [2.3–8.0] | Ref | 5.8 [3.4–9.7] | Ref |
| 25 | 5.0 [3.3–7.5] | 1.15 [0.91–1.40] | 7.5 [5.2–10.5] | 1.31 [1.10–1.52] |
| 35 | 5.8 [4.6–7.4] | 1.37 [0.78–1.97] | 10.2 [8.4–12.2] | 1.84 [1.27–2.41] |
| 45 | 6.1 [4.6–8.1] | 1.46 [0.60–2.32] | 11.8 [9.7–14.2] | 2.19 [1.30–3.08] |
| 55 | 5.3 [3.9–7.2] | 1.25 [0.37–2.14] | 10.5 [8.6–12.7] | 1.91 [1.03–2.79] |
| 65 | 3.7 [2.6–5.1] | 0.85 [0.10–1.60] | 6.8 [5.5–8.4] | 1.19 [0.55–1.83] |
| 75 | 2.2 [1.2–3.8] | 0.50 [0.00–1.19] | 3.6 [2.3–5.5] | 0.60 [0.04–1.17] |
| Ethnicity | | | | |
| White | 4.6 [3.6–5.5] | Ref | 7.7 [6.5–9.0] | Ref |
| Minority ethnic group | 5.6 [2.4–8.8] | 1.09 [0.54–2.23] | 11.0 [7.3–14.7] | 1.37 [0.90–2.10] |
| Occupational social grade | | | | |
| ABC1 (more advantaged) | 5.2 [3.8–6.5] | Ref | 7.5 [6.2–8.8] | Ref |
| C2DE (less advantaged) | 4.1 [2.7–5.6] | 0.81 [0.50–1.31] | 9.2 [7.0–11.3] | 1.32 [0.96–1.82] |
| Financial situation | | | | |
| Living comfortably | 2.9 [1.6–4.3] | Ref | 5.9 [4.0–7.8] | Ref |
| Doing alright | 4.5 [2.9–6.1] | 1.45 [0.81–2.62] | 6.7 [4.9–8.4] | 1.10 [0.70–1.73] |
| Just about getting by | 5.9 [3.8–8.0] | 1.99 [1.10–3.60] | 10.8 [8.2–13.5] | 1.85 [1.20–2.84] |
| Finding it quite difficult | 6.7 [2.6–10.7] | 2.11 [0.87–5.14] | 11.5 [7.1–16.0] | 1.86 [1.07–3.23] |
| Finding it very difficult | 7.7 [2.0–13.4] | 2.41 [0.90–6.47] | 10.2 [4.5–15.9] | 1.59 [0.78–3.22] |
| Economically inactive due to long-term illness or disability | | | | |
| No | 4.3 [3.4–5.3] | Ref | 8.0 [6.7–9.2] | Ref |
| Yes | 12.7 [5.1–20.4] | 2.91 [1.33–6.35] | 11.9 [6.7–17.0] | 1.26 [0.74–2.16] |
| Smoking status | | | | |
| Never | 4.3 [3.0–5.6] | Ref | 7.4 [5.9–8.8] | Ref |
| Former | 4.6 [2.8–6.3] | 1.13 [0.66–1.91] | 10.0 [7.5–12.6] | 1.45 [1.00–2.12] |
| Current | 6.0 [3.4–8.5] | 1.31 [0.73–2.36] | 8.1 [4.8–11.4] | 1.04 [0.65–1.67] |
| Level of alcohol consumption (AUDIT-C)[4] | | | | |
| 0 | 5.4 [3.6–7.9] | Ref | 9.0 [7.1–11.2] | Ref |
| 3 | 4.3 [3.2–5.8] | 0.81 [0.35–1.27] | 6.7 [5.2–8.4] | 0.71 [0.36–1.06] |
| 5 | 4.2 [3.1–5.8] | 0.78 [0.25–1.31] | 7.3 [5.6–9.5] | 0.76 [0.36–1.17] |
| 8 | 4.6 [3.4–6.4] | 0.83 [0.30–1.36] | 11.8 [8.4–16.3] | 1.25 [0.76–1.73] |
| 11 | 5.4 [2.7–10.4] | 0.94 [0.17–1.71] | 20.7 [11.2–35.0] | 2.33 [1.52–3.15] |

*Table continues on next page.*

**Table S3.** *continued*

| | Used GLP-1RAs to support weight loss in the past year[1] | | | |
|---|---|---|---|---|
| | Men | | Women | |
| | % [95% CI] | OR [95% CI][2] | % [95% CI] | OR [95% CI][2] |
| Past-month psychological distress | | | | |
| None/low | 3.1 [2.1–4.0] | Ref | 7.0 [5.6–8.5] | Ref |
| Moderate/severe | 7.9 [5.9–9.8] | 2.62 [1.69–4.07] | 9.8 [7.7–11.9] | 1.33 [0.95–1.85] |
| History of eating disorders[5] | | | | |
| No | 4.7 [3.1–6.3] | Ref | 8.8 [6.6–11.1] | Ref |
| Yes | 7.4 [0.0–25.7] | 1.23 [0.09–16.91] | 12.2 [3.0–21.5] | 1.32 [0.50–3.50] |

CI, confidence interval. OR, odds ratio.

[1] Among adults in Great Britain who had not used a GLP-1RA or other medication for weight loss in the past year.

[2] Adjusted for age.

[3] Predicted estimates from logistic regression models with age modelled using restricted cubic splines. Note that the models used to derive these estimates included data from participants of all ages, not only those who were aged exactly 16, 25, 35, 45, 55, or 65 years.

[4] Predicted estimates from logistic regression models with AUDIT-C score modelled using restricted cubic splines. Note that the models used to derive these estimates included data from all participants who provided data on AUDIT-C, not only those who scored exactly 0, 3, 5, 8, or 11.

[5] History of eating disorders was only assessed in February 2025; analyses were restricted to participants surveyed in this wave.
